# Evaluating the impact of Pharmacare on youth with mental health needs: a regression discontinuity analysis of unmet health care and life stress

**DOI:** 10.64898/2026.03.11.26348184

**Authors:** Peiya Cao, Yihong Bai, Kristine Ienciu, Gwen Ehi, Antony Chum

**Author notes:** Corresponding author: Antony Chum; address: 301F Stong College, York University, 4700 Keele St, Toronto, ON, Canada, M3J 1P3. These authors have contributed equally to this work and share first authorship.

## Abstract

**Background:** Canada’s youth are facing mental health crises due to barriers in accessing timely and affordable care. Ontario’s OHIP+ pharmacare program, introduced in 2018, provided free prescription drug coverage to individuals under 25. While OHIP+ increased prescription use, its effects on perceived access and well-being among youth with mental health needs remain unclear.

**Methods:** We used a regression discontinuity design leveraging the OHIP+ age-eligibility cutoff at 25 to estimate its causal impact on unmet health care needs and self-perceived life stress. The sample included Ontario respondents aged 20–29 (n=1,053) from the 2018–2019 Canadian Community Health Survey who reported needing mental health support. Outcomes were self-reported unmet health care needs and a 5-point life stress scale. Models adjusted for sociodemographic factors and used age in months as the running variable.

**Results:** Loss of OHIP+ eligibility at age 25 was associated with a 19.0 percentage-point increase in the probability of reporting unmet health care needs (95% CI: 0.5 to 37.6 percentage points) and a 1.33-point increase in perceived life stress (95% CI: 0.45 to 2.21). These effects were consistent across subgroups and robust to multiple sensitivity tests.

**Conclusion:** OHIP+ improved access and reduced stress among youth with mental health needs while coverage was in place. However, the abrupt loss of eligibility at age 25 increased unmet needs and psychological strain, underscoring the need for continuous, inclusive pharmacare to support youth well-being.

**What is already known on this topic:** Public drug coverage programs such as OHIP+ have been shown to increase prescription drug use among youth in Ontario, Canada. However, little is known about whether such programs improve patient-perceived outcomes. Existing evaluations have largely relied on descriptive data or aggregate prescribing trends, with few studies examining individual-level outcomes using quasi-experimental methods.

**What this study adds:** Using a regression discontinuity design, this study provides the causal evidence that the OHIP+ pharmacare program reduced unmet health care needs and life stress among youth with perceived mental health concerns while they remained eligible for coverage. The findings show that the abrupt loss of eligibility at age 25 was associated with increased unmet needs and heightened stress, suggesting that age-based cutoffs in drug coverage may disrupt care and contribute to psychological strain during a critical life stage.

**How this study might affect research, practice or policy:** This study suggests that universal drug coverage programs should ensure continuity through young adulthood to avoid worsening access gaps. The findings support the need for a national pharmacare framework that promotes equitable and sustained health support.

## Introduction

Canada is facing a deepening youth mental health crisis. According to the Mental Health Commission of Canada, approximately 1.2 million children and youth are grappling with mental health disorders[1]. Young people aged 15 to 24 are now more likely than any other age group to experience mental health challenges [2]. In Ontario, nearly 40% of high school and post-secondary students are experiencing moderate-to-serious levels of psychological distress[3], underscoring the widespread nature of the crisis. Despite the growing need, many young people continue to face barriers to access timely and appropriate mental health care [4]. In Ontario, the proportion of individuals aged 12 and older who reported an unmet or only partially met mental health care need in the past year was higher than the national average (46.1% versus 43.8%) [5]. These access gaps are especially pronounced among youth aware of their mental health needs but unable to access care. Those transitioning into the adult mental health system are particularly at risk, with less than 20% receiving appropriate treatment [6], a rate that reflects the low treatment rates among children with mental health needs. These youth often remain undiagnosed and unsupported, highlighting critical shortcomings in the current healthcare system.

Addressing these unmet needs requires a multifaceted approach to treatment, often involving a combination of supports—such as counselling, community services, and, when appropriate, medication. However, barriers to accessing these services remain deeply entrenched. Canada is the only country with universal medical health care that does not offer universal pharmacare, leaving many individuals, particularly youth, without coverage for essential treatments. Approximately 12% of Canadians under age 25 lack both public and private prescription drug coverage, and young adults are more likely to face cost-related non-adherence, meaning they delay, skip, or forgo filling prescriptions due to financial constraints [7]. This inequity is further exacerbated by the fragmented structure of provincial drug plans, which vary in eligibility and coverage. For youth without prescription coverage—many of whom are unemployed or in school—these inconsistencies can create delays in care, worsening symptoms, increased suicide risk, and hindered recovery.

Recognizing these challenges, the Ontario government introduced the Ontario Health Insurance Plan Plus (OHIP+) in 2018. This pharmacare program provides free access to over 5,000 prescription medications for residents under the age of 25, however, in April 2019, the program was modified to exclude individuals with access to private insurance or other drug benefits, such as coverage through a parent’s plan or a university/college health plan [8]. OHIP+ offers a unique policy window to evaluate whether expanding drug coverage reduces unmet mental health needs and improves overall well-being among youth. While a growing number of studies have explored the impacts of pharmacare programs on youth in Canada, the overall evidence base remains limited. Several studies have found that OHIP+ increased prescription drug use among children and youth [9–11]. For example, publicly covered prescriptions among youth increased by approximately 290% following implementation, while dispensing of benzodiazepines and prescription stimulants rose by 12.8% and 14.2%, respectively, with the largest increases observed among those aged 20–24. Evidence from Quebec’s 1997 mandatory drug plan similarly showed a 13% increase in medication use and a 10% increase in GP visits, without increases in specialist care or hospital admissions [12]. In contrast, a population-based ARIMA analysis of youth aged 18 and under found no significant changes in antidepressant or antipsychotic dispensing following OHIP+ implementation or modification, though its single-group design and aggregate data limited causal inference and individual-level analysis [13]. Additionally, there is evidence that OHIP+ has improved the glycemic management of low-SES youth with diabetes (measured by HbA1c), relative to their high-SES counterparts [14], with the assumption that those who are low-SES are less likely to be covered by private coverage and least able to afford medications out-of-pocket.

While prior research has shown that OHIP+ increased prescription drug utilization, to our knowledge, no studies have yet examined whether the policy improved perceived access to care and life stress. One key indicator of access is unmet health care need—a patient-reported measure that captures perceived delays or failures in receiving timely and appropriate care. This outcome is particularly salient for youth with mental health challenges, who often recognize their need for help but are unable to access services due to cost, stigma, or inadequate system navigation support. Unmet need reflects more than service availability; it signals the cumulative effect of financial, structural, and psychological barriers. Accordingly, we examine overall unmet health care need among youth and young adults who report a perceived need for mental health–related care, focusing on access to health services conditional on self-identified mental health need. Because OHIP+ aimed to eliminate one of the most tangible obstacles—out-of-pocket costs for medications—it provides an opportunity to assess whether addressing financial barriers translates into fewer instances where young people feel their health care needs go unfulfilled. In this way, unmet health care need serves as a proximate indicator of whether pharmacare can meaningfully improve access to treatment and reduce the burden of untreated mental health concerns among youth. Unlike prescription utilization measures, which capture realized care among those who successfully obtain medications, unmet health care need captures situations in which individuals perceive a need for care but are unable to access it at all, thereby reflecting access barriers that utilization data cannot observe. Life stress is another important outcome, capturing the chronic mental and emotional strain that can negatively impact youth well-being, increase psychological distress, and contribute to maladaptive coping behaviours [15]. Among young people, life stress is often exacerbated by financial insecurity and difficulties managing chronic or emerging health conditions. By improving access to necessary prescription medications, OHIP+ may help reduce this stress through two key mechanisms: first, by supporting more effective management of physical and mental health symptoms; and second, by easing the financial pressures associated with medication costs.

Most existing studies focus on descriptive trends or utilization rates, offering limited insight into causal effects. In contrast, this study employs a regression discontinuity design, leveraging the sharp eligibility cutoff at age 25 to estimate the causal impact of pharmacare in Ontario. This quasi-experimental approach strengthens causal inference by comparing individuals just below and above the age threshold. Our analyses focus on a subset of youth who report a perceived need for mental health–related care, allowing us to examine access barriers among those for whom pharmacare coverage is most directly relevant. Importantly, our sample captures a critical and underrepresented group: youth who are aware of their mental health needs but face significant barriers to accessing formal care. While some may have limited or informal support, they remain underserved, offering insights into how policy can better reach those most in need.

## Methods

### Data source and study population

This study used data from the 2018- 2019 Canadian Community Health Survey (CCHS). The CCHS is a cross-sectional national survey administered by Statistics Canada and Health Canada that provides detailed information about the health status, health determinants, birth dates, and other demographic characteristics of the Canadian population aged 12 years and older [16,17]. For this analysis, we focused on respondents aged 20–29 residing in Ontario who reported a perceived need for mental health–related care during the past 12 months (unweighted N=1,053). The analytic sample was identified using the Canadian Community Health Survey (CCHS) Perceived Need for Care module, which first asked whether respondents had received any help for problems related to emotions, mental health, or substance use, with response options including information, medication, counselling, other help, or none. Respondents who reported receiving any help were asked whether they received as much help as needed, while those who reported receiving no help were asked whether they believed they needed any of these types of help; those answering “yes” for medication, information, or counselling through either pathway were classified as having a perceived need for care and included in the analytic sample. Respondents who reported needing only “other help,” as well as those with missing, “don’t know,” “refusal,” or “not stated” responses, were excluded, and although all included respondents reported a perceived need for care, this does not imply that all had unmet needs.

### Outcomes

The two primary outcomes were: (1) unmet need for health care, and (2) life stress. Unmet need was determined from the responses to questions about the perceived need for health care: “During the past 12 months, was there ever a time when you felt that you needed health care, other than homecare services, but you did not receive it?” Respondents who indicated a health care need were classified into two categories: (a) need met (received sufficient health care), and (b) need not met (received insufficient health care). Unmet health care need was coded as “1” for responses of “b”, indicating an unmet need for health care. In contrast, “0” was assigned to responses of “a,” representing a fulfilled need for health care. Mental health needs often co-occur with broader health service needs [18]; therefore, examining overall unmet health care need among youth and young adults who report a perceived need for mental health care provides a meaningful measure of access to health services in this population. Self-perceived life stress was measured using the question: “Thinking about the amount of stress in your life, would you say that most days are: 1) not at all stressful, 2) not very stressful, 3) a bit stressful, 4) quite a bit stressful, or 5) extremely stressful?”. We treated this variable as a continuous outcome in the analysis to retain its full variability and statistical power, with higher scores reflecting greater perceived stress [19–21].

### Statistical analysis

We used the regression discontinuity (RD) to evaluate the sharp change in unmet medication need for mental health problems and general mental health status at the age cutoff of 25 at the interview in Ontario, according to the OHIP+ policy. By exploiting the exogenous assignment of the OHIP+ policy based on a strict age threshold, the RD approach creates a quasi-experimental setting where individuals just below and above the cutoff are assumed to be similar in both observed and unobserved characteristics. This local randomization minimizes potential confounding and selection biases, thereby enhancing the internal validity of our causal estimates [22]. We employed age in months as the running variable to precisely capture respondents’ ages, calculating each respondent’s age using the provided birth month and interview month from the surveys. Demographic characteristics, including sex, high-school diploma education attainment, ethnicity (white vs. non-white), household income (continuous), employment status and immigration status (immigrants vs. non-immigrants), were included as control variables. We used a user-written package for RD [23], which employs optimal bandwidth selection methods to minimize mean squared error for point estimation and optimize coverage error rate for confidence intervals and applies a triangular kernel weighting that assigns linear down-weighting to the same observations. Following current best practice for RD designs, we report local linear specifications as our main results, because they provide the standard mean-squared-error–optimal estimator at the cutoff and a transparent approximation to the outcome–age relationship near the threshold [23]. We use the single MSE-optimal bandwidth for the main analyses to ensure a common, interpretable comparison window on both sides of the cutoff.

We conducted several supplementary analyses by rerunning the RD model. Since the sample included individuals who reported needing help for mental health in the form of medication, information, or counselling, we performed two sub-sample analyses: (1) those who reported a need for medication, and (2) those who reported a need for information or counselling. Conceptually, this distinction allows us to examine whether changes in medication access may have indirect effects on perceived needs for psychosocial support, such as information or counselling.

Furthermore, we conducted extensive robustness and sensitivity analyses. These included standardizing life stress; re-estimating models using alternative bandwidths, polynomial specifications, and uniform kernel weights; and estimating unadjusted models without covariates. We also examined extra binary outcomes, self-perceived poor mental health [24] and poor general health [25], where “fair” and “poor” are considered as “poor” [26,27]; Placebo and falsification tests were conducted using a pre-policy year, alternative age cutoffs, and respondents from other provinces. Additional analyses assessed the April 2019 OHIP+ modification, donut RD models addressing variation in coverage windows, and alternative analytic samples. All analyses were conducted using STATA v18, with two-tailed tests and a 5% significance level.

## Results

Table 1 presents descriptive characteristics of Ontario respondents aged 20–29 from the 2018 and 2019 Canadian Community Health Survey, stratified by age group (20–24 vs. 25–29). In both years, individuals aged 25–29 accounted for a larger proportion of the sample. In 2019, unmet health care needs were more common among those aged 25–29 (15.5%) than 20–24 (10.1%). Older respondents were more likely to be male, white, employed, and have slightly lower household income. Immigrant representation was higher among the younger group. In 2018, some similar patterns were observed. The 25–29 group reported comparable life stress but had higher income and educational attainment, and were less likely to be immigrants.

**Table 1.** Characteristics of Ontario youths aged 20–29 with mental health needs.

|  | Age 20-24 | Age 25-29 |
| --- | --- | --- |
| CCHS 2019 |  |  |
| Proportion of the sample | 65.6% | 34.4% |
| Unmet health care needs | 10.1% | 15.5% |
| Age (mean/sd) | 22.3 (1.5) | 27.4 (1.2) |
| Male | 53.2% | 64.1% |
| White | 55.0% | 74.8% |
| Household income (per 1000 dollars) | 132.9 (7.7) | 125.0 (9.8) |
| High-school education | 91.6% | 90.1% |
| Employed | 84.6% | 87.9% |
| Immigrants | 19.1% | 14.4% |
| CCHS 2018 |  |  |
| Proportion of the sample | 64.4% | 35.6% |
| Life stress (mean/sd) | 3.3 (0.8) | 3.3 (0.8) |
| Age (mean/sd) | 22.7 (1.6) | 27.4 (0.9) |
| Male | 61.8% | 64.0% |
| White | 67.0% | 86.9% |
| Household income (per 1000 dollars) | 93.7 (4.4) | 118.6 (5.1) |
| High-school education | 93.4% | 96.4% |
| Employed | 90.3% | 87.2% |
| Immigrants | 23.9% | 10.2% |

Table 2 presents the linear regression discontinuity estimates of the impact of the OHIP+ pharmacare policy on unmet health care needs and self-perceived life stress. At the age-25 cutoff, the implementation of OHIP+ resulted in a significant 19.0 percentage-point increase in the probability of reporting unmet health care needs (95% CI: 0.5 to 37.6 percentage points). The policy was also associated with a significant increase in self-perceived life stress score (β = 1.333, 95% CI: 0.454 to 2.212), where higher scores indicate greater perceived stress. These findings are visually depicted in Figure 1. Panel A and Panel B illustrate the regression discontinuity plots for unmet health care needs and life stress, respectively. Both panels show clear upward shifts in the predicted outcomes at the age-25 cutoff, consistent with the estimates in Table 2.

**Figure 1a:**
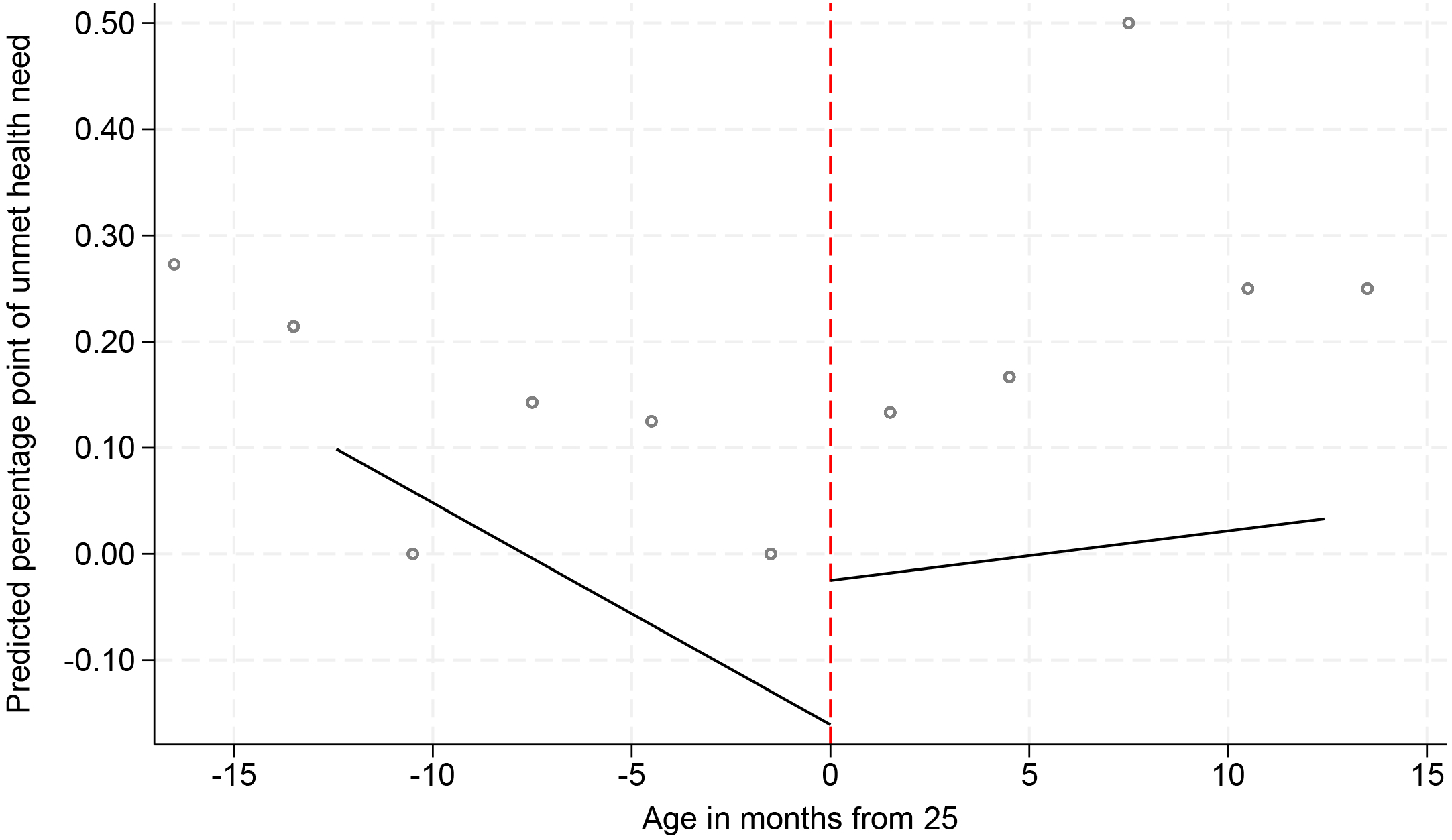
Discontinuity in perceived unmet health need at age 25.

**Figure 1b:**
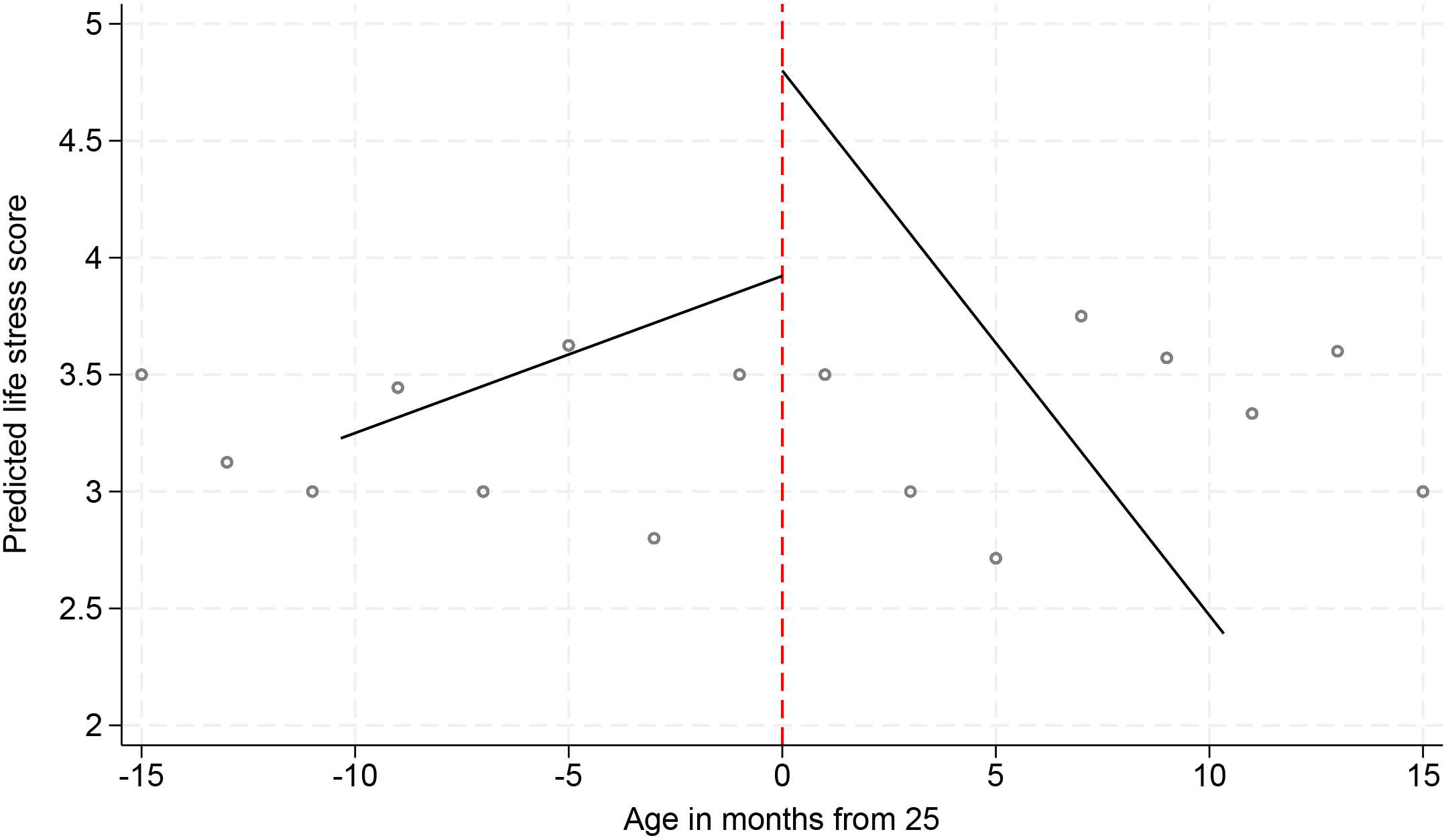
Discontinuity in perceived life stress at age 25.

**Table 2.**
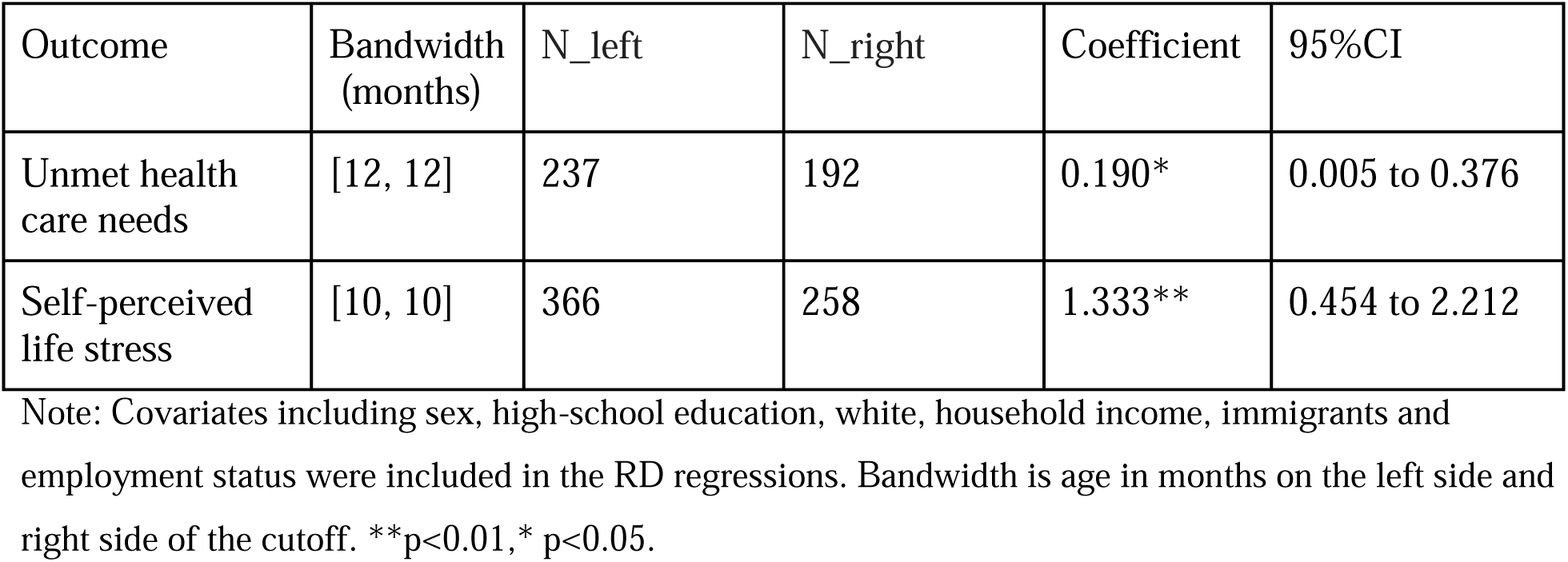
Linear regression discontinuity estimates of the impact of OHIP+ on self-rated unmet health care needs and life stress.

Robustness checks consistently supported the main findings. First, results for life stress were robust to z-score standardization (Table S1). Second, estimates were stable across alternative bandwidths and polynomial specifications (Table S2), with unmet health care needs increasing by approximately 19–20 percentage points at the age-25 cutoff (p<0.05) and self-perceived life stress increasing by 1.35–1.69 points on the 5-point scale (p<0.01 for linear; p<0.05 for quadratic). Third, results were similar when using fixed bandwidths of 12 and 24 months (Table S3), unadjusted models without covariates (Table S4), and uniform kernel weighting (Table S5). Fourth, subgroup analyses showed increased unmet health care needs among individuals reporting a need for medication only as well as information and counselling only (Table S6).

Analyses of supplementary outcomes showed no statistically significant discontinuities in self-perceived poor mental health or poor general health (Table S7). Placebo and falsification tests—including a non-policy year (2017; Table S8), alternative age cutoffs at ages 24 and 26 (Table S9), and respondents from other provinces without OHIP+ (Table S10)—yielded non-significant estimates. Balance tests confirmed no discontinuities in baseline covariates at the age-25 cutoff (Table S11). No significant effects were observed for the April 2019 OHIP+ policy change (Table S12). Donut RD models excluding individuals within ±6 months of age 25 produced similar results (Table S13). Additional analyses showed attenuated effects in the full Ontario sample (Table S14) and no discontinuity in the probability of reporting a mental health care need at the cutoff (Table S15).

## Discussion

This study evaluated the impact of Ontario’s OHIP+ pharmacare program on unmet health care needs and self-perceived life stress among youth with mental health concerns, using a regression discontinuity design. Our results showed a significant increase in the probability of reporting unmet health care needs and higher self-perceived stress levels at the age-25 eligibility cutoff. These findings remained consistent across subgroups and were robust to multiple sensitivity analyses, including placebo tests and falsified policy years.

One possible explanation for the increase in unmet need is that the transition out of OHIP+ at age 25 introduces abrupt disruptions in care, particularly for youth who had relied on free access to medications. As individuals lose coverage, they may experience difficulty affording prescriptions, navigating private plans, or re-entering the healthcare system as adults [28]. The observed increase in unmet needs suggests that losing OHIP+ coverage at age 25 disrupts access to care, particularly for youth with chronic or recurring mental health issues who had relied on free medications. Importantly, this pattern does not necessarily imply immediate deterioration in overall mental health status, but rather reflects changes in perceived access and care experiences following the loss of coverage.

Similarly, the observed rise in life stress suggests that the expiration of OHIP+ may introduce psychological and financial burdens. While pharmacare policies are designed to reduce cost-related barriers to medication [29,30], the age-based cutoff creates a sharp transition that may lead to treatment discontinuity. Youth nearing age 25 often face additional stressors—such as unstable employment [31], student loan debt [32], or housing precarity [33] —which may compound the impact of losing pharmacare support. The absence of short-term changes in self-perceived mental or general health suggests that the RD estimates are capturing proximal stress responses to coverage loss, rather than downstream changes in health outcomes. While the research on OHIP+ is still scarce, there have been a few studies that investigated the impact of Medicare Part D on well-being and found a similar effect. For example, one study found that Medicare Part D significantly reduced depressive symptoms among older adults [34], and individuals with continuous enrollment were less likely to have worse self-rated health [35].

While previous studies have documented increased prescription volumes following OHIP+ implementation [9,10], few have investigated how such policies affect patient-perceived outcomes. Moreover, most existing analyses rely on aggregate dispensing data or descriptive trends, limiting their ability to draw causal conclusions. By contrast, our use of a regression discontinuity design and individual-level survey data allows for a more nuanced assessment of how coverage policies shape youth experiences at a critical life stage. Our results highlight the importance of examining how youth feel about their ability to access care and manage mental health demands, particularly as they age out of public support.

From a policy perspective, our findings offer evidence that age-based cutoffs in pharmacare programs, such as OHIP+, may unintentionally disrupt care and contribute to greater unmet health needs and life stress among youth. As Canada considers the implementation of a national pharmacare program, this study provides support for more continuous and inclusive coverage (such as a universal pharmacare system), particularly for young adults navigating mental health challenges, financial precarity, and transitions out of public support. While drug coverage alone is not a complete solution, these results suggest that eliminating cost-related barriers to medications can play a critical role in improving access and well-being, especially when paired with other supports such as counselling and system navigation.

This study has several limitations. First, the CCHS is cross-sectional, and although the regression discontinuity design strengthens causal inference, we cannot track individuals over time to assess longer-term impacts. Second, the outcomes—unmet need and life stress—are self-reported and subject to recall or reporting bias. Third, we are unable to directly measure medication adherence, dosage, or specific mental health diagnoses, which may influence the observed effects. Fourth, while our findings provide strong internal validity for youth around the age-25 cutoff in Ontario, generalizability to other age groups, provinces, or international settings may be limited due to differences in health system structures, coverage policies, and population characteristics. Nonetheless, the study’s strengths include the use of individual-level data, a strong quasi-experimental design, and multiple robustness checks to ensure internal validity. Fifth, analytic sample sizes were relatively small, resulting in wide confidence intervals. Future analyses using larger datasets could improve statistical power. However, our findings were consistent across multiple robustness checks using alternative model specifications and estimation strategies.

## Conclusion

In conclusion, the abrupt increase in unmet needs and heightened life stress at the age-25 cutoff suggests that OHIP+ effectively reduced barriers to care and supported well-being among youth with perceived mental health needs while they remained eligible for the program. These results suggest that the program helped reduce barriers to care and supported youth well-being while in place, the abrupt loss of coverage may disrupt treatment and introduce new psychological and financial burdens during a critical transition period. As Canada explores a national pharmacare strategy, our findings provide evidence supporting the value of continuous and inclusive drug coverage—particularly for young adults facing mental health challenges—to promote sustained access and improved health outcomes.

## Declarations of competing interest

None.

## Acknowledgements

This study is based on data provided by Statistics Canada through the Canadian Community Health Survey (CCHS). The authors thank Statistics Canada for access to the data and support in facilitating this research. The opinions expressed do not represent the views of Statistics Canada. The authors also thank Qian Liu at Brock University for valuable suggestions.

## Contributors

Peiya Cao: Conceptualization, Formal analysis, Investigation, Methodology, Resources, Software, Validation, Writing - Original Draft, Writing - review & editing

Yihong Bai: Conceptualization, Investigation, Methodology, Resources, Software, Validation, Writing - Original Draft, Writing - review & editing

Kristine Ienciu: Investigation, Writing - review & editing

Gwen Ehi: Writing - review & editing

Antony Chum: Funding acquisition, Investigation, Methodology, Project administration, Resources, Supervision, Validation, Writing - Original Draft, Writing - review & editing

Peiya Cao and Yihong Bai shared the first authorship.

## Funding

This project was funded by the Canadian Institutes of Health Research (Project Grant FRN# 525784, NPI: Antony Chum). The project principal investigator (Antony Chum) is supported by the Canada Research Chair Program (grant CRC-2021-00269). This project is also supported by the Karen Maxwell Foundation (no grant number applicable).

## Ethics approval

Ethics approval for this study was obtained through York University (REB#-2025-226).

## Competing interests

None.

## Patient consent for publication

Not applicable.

## Data Availability Statement

This study utilized de-identified microdata accessed through the Research Data Centre (RDC) program managed by Statistics Canada. The data are protected by strict confidentiality provisions under the Statistics Act and are accessible only to researchers with approved projects who meet security and confidentiality requirements. The data are not publicly available but can be accessed upon request to Statistics Canada through their RDC program, subject to similar security, confidentiality, and ethical requirements.

## Supplementary material

**Table S1.** Linear regression discontinuity estimates of the impact of OHIP+ on self-perceived life stress (z-score)

| Outcome | Bandwidth (months) | N_left | N_right | Coefficient | 95% CI |
| --- | --- | --- | --- | --- | --- |
| Self-perceived life stress (z-score) | [10, 10] | 366 | 258 | 1.480** | 0.504 to 2.457 |

Note: Estimates are from fully adjusted regression discontinuity models controlling for sex, high-school education, white, household income, immigrants and employment status. The single mean squared error (MSE)–optimal bandwidths are used. \*\*p<0.01.

**Table S2.** Regression discontinuity estimates of the impact of OHIP+ on self-rated unmet health care needs and life stress under alternative bandwidths, linear and quadratic trends.

| Outcome | Order of local polynomial | Bandwidth (months) | N_left | N_right | Coefficient | 95% CI |
| --- | --- | --- | --- | --- | --- | --- |
| Unmet health care needs | Linear | [13, 15] | 237 | 192 | 0.193* | 0.014 to 0.371 |
|  | Quadratic | [19, 21] |  |  | 0.204* | 0.002 to 0.407 |
| Self-perceived life stress | Linear | [15, 6] | 366 | 258 | 1.348** | 0.347 to 2.349 |
|  | Quadratic | [27, 10] |  |  | 1.693** | 0.487 to 2.899 |

Note: Estimates are from fully adjusted regression discontinuity models controlling for sex, high-school education, white, household income, immigrants and employment status. Two-sided MSE-optimal bandwidths were used. \*\* $p < 0.01$ , \* $p < 0.05$ .

**Table S3.** Regression discontinuity results for unmet health care needs and life stress with fixed bandwidths (12 and 24 months)

| Outcome | Order of local polynomial | Bandwidth (months) | N_left | N_right | Coefficient | 95%CI |
| --- | --- | --- | --- | --- | --- | --- |
| Unmet health care needs | Linear | [12, 12] | 237 | 192 | 0.195* | 0.007 to 0.384 |
|  |  | [24, 24] |  |  | 0.190* | 0.006 to 0.375 |
|  | Quadratic | [12, 12] |  |  | 0.297* | 0.040 to 0.553 |
|  |  | [24, 24] |  |  | 0.211* | 0.018 to 0.403 |
| Self-perceived life stress | Linear | [12, 12] | 366 | 258 | 1.184** | 0.339 to 2.029 |
|  |  | [24, 24] |  |  | 0.745** | 0.136 to 1.354 |
|  | Quadratic | [12, 12] |  |  | 2.560*** | 1.535 to 3.584 |
|  |  | [24, 24] |  |  | 0.830* | 0.174 to 2.117 |

Note: Estimates are from fully adjusted regression discontinuity models controlling for sex, high-school education, white, household income, immigrants and employment status. The single MSE-optimal bandwidths are used. \*\*\* $p < 0.01$ , \*\* $p < 0.01$ , \* $p < 0.05$ .

**Table S4.** Unadjusted regression discontinuity estimates of the impact of OHIP+ on unmet health care needs and self-perceived life stress.

| Outcome | Order of local polynomial | Bandwidth (months) | N_left | N_right | Coefficient | 95%CI |
| --- | --- | --- | --- | --- | --- | --- |
| Unmet health care needs | Linear | [12, 12] | 348 | 207 | 0.257** | 0.071 to 0.444 |
|  | Quadratic | [15, 15] |  |  | 0.343** | 0.131 to 0.554 |
| Self-perceived life stress | Linear | [8, 8] | 404 | 306 | 1.500** | 0.231 to 2.768 |
|  | Quadratic | [11, 11] |  |  | 2.440** | 0.979 to 3.901 |

Note: The single MSE-optimal bandwidths are used. \*\*p<0.01,\* p<0.05.

**Table S5.** Regression discontinuity estimates of the impact of OHIP+ on unmet health care needs and self-perceived life stress using uniform kernel weighting with linear and quadratic age trends.

| Outcome | Order of local polynomial | Bandwidth (months) | N_left | N_right | Coefficient | 95%CI |
| --- | --- | --- | --- | --- | --- | --- |
| Unmet health care needs | Linear | [12, 12 ] | 237 | 192 | 0.206* | 0.027 to 0.461 |
|  | Quadratic | [19, 19] |  |  | 0.235* | 0.016 to 0.455 |
| Self-perceived life stress | Linear | [8, 8 ] | 366 | 258 | 1.548* | 0.314 to 2.781 |
|  | Quadratic | [13, 13] |  |  | 1.483** | 0.420 to 2.545 |

Note: Estimates are from fully adjusted regression discontinuity models controlling for sex, high-school education, white, household income, immigrants and employment status. The single MSE-optimal bandwidths are used. \*\*p<0.01,\* p<0.05.

**Table S6.** Linear regression discontinuity estimates of the impact of OHIP+ on unmet health care needs among individuals with mental health medication needs only and those with mental health information/counselling needs only.

| Outcome | Bandwidth (months) | N_left | N_right | Coefficient | 95%CI |
| --- | --- | --- | --- | --- | --- |
| Unmet health care need (medication only) | [10, 10] | 102 | 108 | 0.432* | 0.073 to 0.792 |
| Unmet health care need<br>(information &<br>counselling only) | [15, 15] | 487 | 378 | 0.189* | 0.036 to 0.342 |

Note: Estimates are from fully adjusted regression discontinuity models controlling for sex, high-school education, white, household income, immigrants and employment status. The single MSE-optimal bandwidths are used. \* $p < 0.05$ .

**Table S7.** Linear regression discontinuity estimates of the impact of OHIP+ on self-perceived poor mental health and poor general health among individuals with mental health needs.

| Outcome | Bandwidth<br>(months) | N_left | N_right | Coefficient | 95%CI |
| --- | --- | --- | --- | --- | --- |
| Self-perceived poor mental<br>health | [12, 12] | 366 | 258 | 0.217 | -0.039 to 0.472 |
| Self-perceived poor health | [8, 8] |  |  | -0.084 | -0.331 to 0.162 |

Note: Estimates are from fully adjusted regression discontinuity models controlling for sex, high-school education, white, household income, immigrants and employment status. The single MSE-optimal bandwidths are used.

**Table S8.** Linear regression discontinuity estimates of unmet health care needs and life stress using a falsified policy year of 2017.

| Outcome | Bandwidth (months) | N_left | N_right | Coefficient | 95%CI |
| --- | --- | --- | --- | --- | --- |
| Unmet health care needs | [14, 14] | 404 | 306 | 0.455 | -0.054 to 0.963 |
| Self-perceived life stress | [12,12] | 634 | 792 | -0.180 | -1.280 to 0.920 |

Note: Estimates are from fully adjusted regression discontinuity models controlling for sex, high-school education, white, household income, immigrants and employment status. The single MSE-optimal bandwidths are used.

**Table S9.** Linear regression discontinuity results on the impact of OHIP+ on unmet health care needs and life stress using falsified age cutoffs.

| Age cutoff | Outcome | Bandwidth( months) | N_left | N_right | Coefficient | 95%CI |
| --- | --- | --- | --- | --- | --- | --- |
| 24 years (288 months) | Unmet health care needs | [16, 16] | 237 | 192 | -0.070 | -0.383 to 0.243 |
|  | Self-perceived life stress | [11, 11] | 366 | 258 | 0.527 | -0.345 to 1.398 |
| 26 years<br>(312<br>months) | Unmet health<br>care needs | [10, 10] | 237 | 192 | 0.252 | -0.398 to<br>0.715 |
|  | Self-perceived<br>life stress | [15, 15] | 366 | 258 | 0.675 | -1.100 to<br>2.449 |

Note: Estimates are from fully adjusted regression discontinuity models controlling for sex, high-school education, white, household income, immigrants and employment status. The single MSE-optimal bandwidths are used.

**Table S10.** Linear regression discontinuity estimates of unmet health care needs and life stress in 2018 among respondents from all provinces except Ontario, Quebec, and British Columbia.

| Outcome | Bandwidth | N_left | N_right | Coefficient | 95%CI |
| --- | --- | --- | --- | --- | --- |
| Unmet health<br>care needs | [18, 18] | 181 | 251 | 0.192 | -0.003 to<br>0.333 |
| Self-perceived<br>life stress | [18, 18] | 167 | 211 | -0.292 | -1.025 to<br>0.441 |

Note: Estimates are from fully adjusted regression discontinuity models controlling for sex, high-school education, white, household income, immigrants and employment status. The single MSE-optimal bandwidths are used.

**Table S11.** Regression discontinuity estimates of the impact of turning age 25 on baseline covariates.

| Variables | Coefficient | 95% CI |
| --- | --- | --- |
| Male | -0.134 | -0.904 to 0.635 |
| Household income | 43726 | -17241.9 to 104694.0 |
| High-school education | -0.006 | -0.326 to 0.314 |
| Employed | 0.162 | -0.248 to 0.572 |
| White | -0.103 | -0.795 to 0.589 |
| Immigrants | 0.150 | -0.364 to 0.665 |

**Table S12.** Regression discontinuity estimates of the impact of the OHIP+ amendment on self-rated unmet health care needs and life stress, using linear and quadratic age trends.

| Outcome | Order of local polynomial | N_left | N_right | Bandwidth | Coefficient | 95%CI |
| --- | --- | --- | --- | --- | --- | --- |
| Unmet health care needs | Linear | 342 | 186 | [13, 13] | -0.204 | -0.467 to 0.060 |
|  | Quadratic |  |  | [16, 16] | -0.176 | -0.546 to 0.194 |
| Self-perceived life stress | Linear | 428 | 254 | [21, 21] | 0.043 | -0.800 to 0.886 |
|  | Quadratic |  |  | [27, 27] | -0.068 | -1.070 to 0.933 |

Note: Unmet health care needs from 2020 CCHS (past 12 months recall covers OHIP+ amendment period starting from April 2019) and self-perceived life stress from 2019 CCHS (interview period includes April 2019 amendment rollout). Estimates are from fully adjusted regression discontinuity models controlling for sex, high-school education, white, household income, immigrants and employment status. The single MSE-optimal bandwidths are used.

**Table S13.** Donut (±6-month) linear regression discontinuity estimates of the impact of OHIP+ on self-perceived mental health and general health among individuals with mental health needs, using fixed bandwidths.

| Outcome | Bandwidth (months) | N_left | N_right | Coefficient | 95%CI |
| --- | --- | --- | --- | --- | --- |
| Unmet health need | [18, 18] | 225 | 159 | 0.843* | 0.009 to 1.677 |
|  | [24, 24] |  |  | 0.698* | 0.038 to 1.357 |
| Self-perceived life stress | [18, 18] | 336 | 242 | 1.840* | 0.403 to 3.277 |
|  | [24, 24] |  |  | 1.574* | 0.345 to 2.803 |

Note: Estimates are from fully adjusted regression discontinuity models controlling for sex, high-school education, white, household income, immigrants and employment status. \* $p < 0.05$ .

**Table S14.** Linear regression discontinuity analyses of the impact of OHIP+ on self-rated unmet health care needs and perceived life stress among Ontarians aged 20–29 years.

| Outcome | Bandwidth (months) | N_left | N_right | Coefficient | 95%CI |
| --- | --- | --- | --- | --- | --- |
| Unmet health care needs | [11, 11] | 799 | 661 | 0.080* | 0.003 to 0.158 |
| Self-perceived life stress | [8, 8] | 1388 | 1082 | 0.966** | 0.390 to 1.542 |

Note: Estimates are from fully adjusted regression discontinuity models controlling for sex, high-school education, white, household income, immigrants and employment status. The single MSE-optimal bandwidths are used. \*\*p<0.01,\* p<0.05.

**Table S15.**
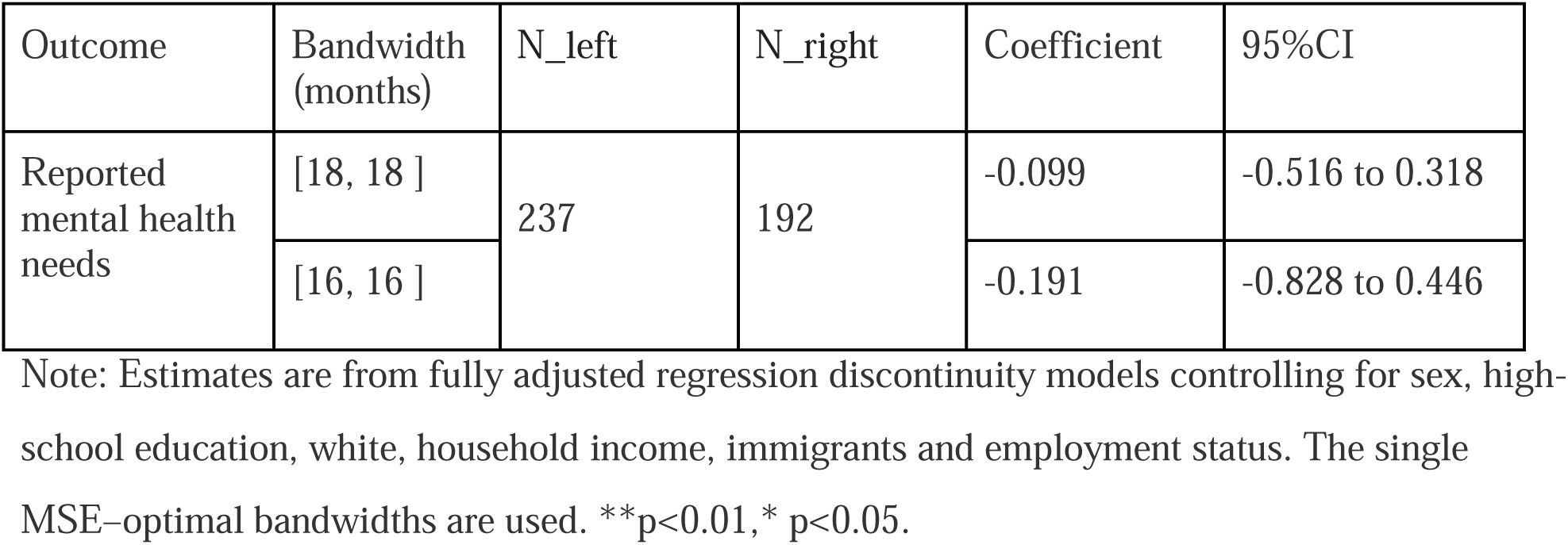
Linear regression discontinuity analyses of reported mental health needs among Ontarians aged 20–29 years.

## Notes

### Competing Interest Statement

The authors have declared no competing interest.

